# A shorter symptom-onset to remdesivir treatment (SORT) interval is associated with a lower mortality in moderate-to-severe COVID-19: A real-world analysis

**DOI:** 10.1101/2020.11.05.20226373

**Authors:** Ravindra M Mehta, Sameer Bansal, Suhitha Bysani, Hariprasad Kalpakam

**Affiliations:** Apollo Super Specialty Hospital, Jayanagar, Bangalore

**Keywords:** COVID-19, earlier initiation, moderate, mortality, remdesivir, safety, severe, SORT

## Abstract

**Background:** Remdesivir is the current recommended anti-viral treatment in moderate-to-severe COVID-19. However, data on several aspects of its use such as impact of timing of therapy, efficacy, and safety in this severity category are limited. The current study aimed to evaluate the impact of timing of remdesivir initiation (symptom-onset to remdesivir treatment [SORT] interval) on in-hospital all-cause mortality in patients with moderate-to-severe COVID-19.

**Methods:** This retrospective study was conducted between June 25, 2020 and October 3, 2020, at a tertiary care dedicated COVID center in India. Consecutive patients with moderate-to-severe COVID-19 (moderate: SpO2 <94%; severe: SpO2 <90%) were included. Data were collected from the health records of the hospital. Remdesivir was administered along with other standard medications as per protocol. The main outcome was the impact of SORT interval on in-hospital all-cause mortality. Subgroups were formed based on SORT interval. Other measures analyzed included overall in-hospital mortality, length of hospital stay, and safety.

**Results:** Of 350 patients treated with remdesivir, 346 were included for the final analysis (males: 270 [78.0%]; median [range] age: 60 [24-94] years). Overall, 243 (70.2%) patients had ≥1 comorbidity; 109 (31.5%) patients had moderate disease, 237 (68.5%) had severe disease, and 50 (14.5%) patients required mechanical ventilation. Of the 346 patients, 76 (22.0%) patients died (moderate: 3 [2.8%], severe: 73 [30.8%]). In the subset of mechanically ventilated patients, 43 (86.0%) died. All-cause mortality was significantly lower in patients with SORT interval ≤9 days (n = 260) compared with those with SORT interval >9 days (n = 86; 18.1% vs 33.7%; *P* = .004). The odds of death were significantly lower in patients with SORT interval ≤9 days vs >9 days (odds ratio = 0.44; 95% CI, 0.25-0.76; *P* = .004). Adverse events (transaminitis ≥5 times upper limit of normal or estimated glomerular filtration rate <30ml/min) leading to drug discontinuation were seen in 4 (1.1%) patients.

**Conclusion:** In this large series of moderate-to-severe COVID-19, initiation of remdesivir ≤9 days from symptom-onset was associated with a significant mortality benefit. These findings indicate a treatment window and reinforce the need for earlier remdesivir initiation in moderate-to-severe COVID-19 infection.

## Introduction

Coronavirus disease 2019 (COVID-19), caused by severe acute respiratory syndrome coronavirus 2 (SARS-CoV-2),^1^ has resulted in more than 1.18 million deaths worldwide as of October 31, 2020.^2^

Remdesivir, a nucleotide analog prodrug with broad antiviral activity, has demonstrated *in vitro* activity against SARS-CoV-2 and *in vivo* activity in a primate model of SARS-CoV-2 infection.^3,4^ Remdesivir received FDA Emergency Use Authorization (EUA) on May 1, 2020 for treatment of moderate-to-severe COVID-19.^5^ On October 22, 2020, the FDA approved remdesivir as the first COVID-19 anti-viral agent in adult and pediatric patients (aged ≥12 years and weighing at least 40 kg) requiring hospitalization.^6^

The efficacy and safety of remdesivir in COVID-19 have been studied in a few clinical trials.^7–10^ In a randomized double-blind trial, hospitalized COVID-19 patients treated with remdesivir had a shorter time to recovery and a trend towards lower mortality at 15 and 29 days compared with placebo.^10^ Research indicates the potential benefits of early initiation of remdesivir in patients with COVID-19. Remdesivir therapy was associated with earlier clinical improvement (18 vs 23 days) and trend towards a lower 28-day mortality (11% vs 15%) in adult patients hospitalized for severe COVID-19 treated within 10 days of symptoms compared with placebo.^7^ However, the recent WHO sponsored Solidarity trial results raised several questions on the efficacy of remdesivir in COVID-19.^11^

Inconsistent data about the efficacy of remdesivir are attributable to various factors including the heterogenous nature of COVID-19 treatment in the pandemic as treatment options evolved, lack of clarity of the optimal timing of initiation of the drug, treatment efficacy assessed from either ‘symptom-onset’ or ‘diagnosis-onset,’ and the difference in ‘standard of care’ across the globe. Specifically, the relationship between timing of remdesivir initiation (from symptom onset) and outcomes has not been clearly elucidated. There is a need to evaluate the effect of optimal remdesivir initiation timing that would improve outcomes in COVID-19, especially in the sicker subsets of moderate-to-severe disease. This study aimed to evaluate the impact of symptom-onset to remdesivir treatment (SORT) interval on clinical outcomes in the subsets of moderate-to-severe COVID-19.

## Methods

### Study Design

This single-center retrospective study was conducted between June 25, 2020 and Oct 3, 2020 at a tertiary dedicated COVID care hospital in adult patients with moderate-to-severe COVID-19 in Bangalore, India. Data on patient demographics, clinical characteristics, and outcomes were collected from health records of the hospital. Ethical approval was obtained from the Institutional Review Board of the participating site. De-identified patient data were used for analysis.

### Study Population

Inclusion criteria were hospitalized patients with SARS-CoV-2 infection as confirmed by reverse-transcription polymerase chain reaction assay, radiologic evidence of pneumonia, and peripheral oxygen saturation (SpO2) of <94% (while breathing room air).

Moderate disease was defined as presence of hypoxia (SpO2 <94%, range: 90%-94%) while breathing room air and respiratory rate ≥24 breaths per minute.^12,13^ Severe disease was defined as presence of clinical signs of pneumonia and one of the following criteria: SpO2 <90% while breathing room air, respiratory rate >30 breaths per minute, heart rate >120 beats per minute, presence of acute respiratory distress syndrome (ARDS), sepsis or septic shock.^12,13^

### Treatment

Patients received intravenous remdesivir 200 mg on day 1 as loading dose, followed by a maintenance dose of 100 mg daily for a total of 5 to 10 days, manufactured by three different pharmaceutical companies. Patients also received other drugs as per protocol, including corticosteroids, anticoagulants, and other supportive therapy. Few patients also received other experimental therapies, including convalescent plasma, tocilizumab, and thrombolytics.

### Outcome Measures

Data on clinical and laboratory findings, including mortality, length of hospital stay (LOHS), and safety outcomes (adverse events [AEs], serious adverse events [SAEs], and suspected drug-related hypersensitivity reactions) were extracted from the medical records.

Demographic parameters analyzed included age, sex, and comorbidities (diabetes mellitus, hypertension, CKD, chronic heart disease [CHD], and chronic respiratory diseases among others). The main outcome measure analyzed was the impact of SORT interval on in-hospital all-cause mortality. Other outcomes analyzed included in-hospital all-cause mortality, AEs, SAEs, treatment-emergent AEs, and overall LOHS.

### Statistical Analysis

Results were summarized descriptively as numbers and percentages, means and standard deviations, or medians and inter-quartile ranges. The difference between means of continuous variables was assessed using t-test/Mann Whitney-U test and difference between proportions of categorical variables, using Fisher’s exact test/Chi-square test.

Kaplan-Meier plots were used to determine the impact of SORT interval (≤6 vs >6, ≤7 vs >7, ≤8 vs >8, ≤9 vs >9, and ≤10 vs >10 days) on survival probability (all-cause mortality) and the differences in the sub-groups were determined using log-rank test. Multivariate regression analysis was carried out to compare mortality outcomes of the cohorts with a specific SORT interval based on information derived from the Kaplan-Meier plots, and odds ratios (ORs) and 95% confidence interval (CI) were calculated.

Differences in outcomes between the study groups were considered significant if *P* <0.05 (two-sided). All statistical calculations were performed using R software (R 4.0.2).

## Results

A total of 350 patients with moderate-to-severe COVID-19 infection received remdesivir, of which 346 were included for the final analysis (4 patients were excluded due to SAEs leading to drug discontinuation). The median (range) age was 60 (24-94) years, with the majority being male (n = 270; 78.0%; **Table 1**). Among the hospitalized patients, one-third had moderate disease (n = 109; 31.5%), and 237 (68.5%) had severe disease. Fifty patients (14.5%) required mechanical ventilation. Most of the patients had ≥1 comorbidity (n = 243; 70.2%), including diabetes mellitus (n = 173; 50.0%), hypertension (n = 163; 47.1%), CHD (n = 54; 15.6%), CKD (n = 18; 5.2%), and chronic respiratory disease (asthma/chronic obstructive pulmonary disease [COPD]; n = 12; 3.5%).

**Table 1.** Demographic and Clinical Characteristics of the Study Population^a^.

| Characteristics | N = 346 |
| --- | --- |
| Age (years), median (range) | 60 (24-94) |
| Male, No. (%) | 270 (78.0) |
| Comorbidities |  |
| Any comorbidity, No. (%) | 243 (70.2) |
| HTN, No. (%) | 163 (47.1) |
| DM, No. (%) | 173 (50.0) |
| CKD, No. (%) | 18 (5.2) |
| CHD, No. (%) | 54 (15.6) |
| Chronic respiratory diseases (asthma, COPD), No. (%) | 12 (3.5) |
| Disease (COVID-19) severity |  |
| Moderate, No. (%) | 109 (31.5) |
| Severe, No. (%) | 237 (68.5) |
| Respiratory support |  |
| Mechanical ventilation, No. (%) | 50 (14.5) |
| Others (NIV/HFNC/oxygen), No. (%) | 296 (85.5) |

Abbreviations: CHD, chronic heart disease; CKD, chronic kidney disease; COPD, chronic obstructive pulmonary disease; COVID-19, coronavirus disease 2019; DM, diabetes mellitus; HFNC, high flow nasal cannula; HTN, hypertension; NIV, non-invasive ventilation.
<sup>a</sup> Of the 350 patients, 4 were excluded from the analysis due to drug discontinuation, and 346 were analyzed.

In addition to remdesivir, all patients received corticosteroids, 131 (37.9%) received convalescent plasma, and 37 (10.7%) received tocilizumab.

Of the 346 admitted patients, 270 (78.0%) were discharged, and 76 (22.0%) died (**Table 2**). In the moderate group, 106 (97.2%) patients were discharged, and 3 (2.8%) died. In the severe group, 164 (69.2%) patients were discharged, and 73 (30.8%) died. In the mechanically ventilated patients, 7 (14.0%) patients were discharged, and 43 (86.0%) died.

**Table 2.**
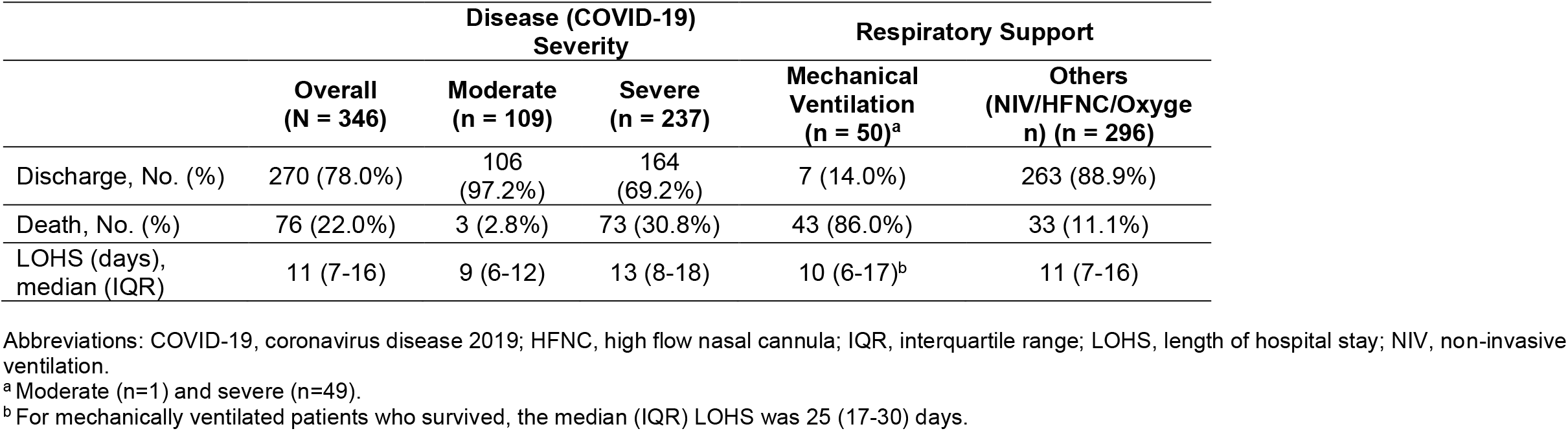
Clinical Outcomes Based on Severity and Respiratory Support.

The median (interquartile range [IQR]) LOHS was 11 (7-16) days in the overall group, 9 (6-12) days in the moderate group, and 13 (8-18) days in the severe group. In mechanically ventilated patients, the overall LOHS was 10 (6-17) days and in those who survived, it was 25 (17-30) days.

Kaplan-Meier plots demonstrated significant difference in the probability of survival for SORT interval ≤9 vs >9 days (*P* = .03; **eFigure 1)** but not for the other intervals. Based on this, the two subgroups of SORT interval ≤9 vs >9 days were further analyzed to evaluate the impact of the timing of remdesivir initiation on outcomes.

**Figure 1.**
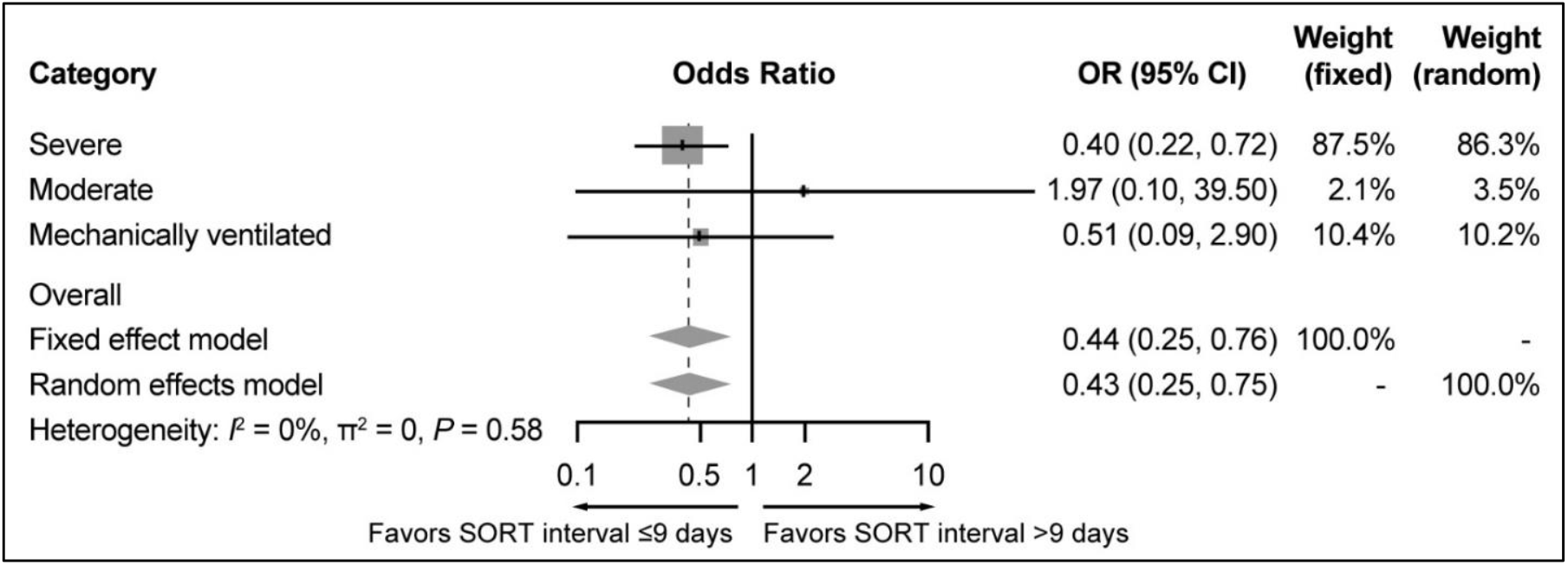
Forest Plot for Mortality Outcome Comparing SORT Interval ≤9 days vs >9 days. Abbreviations: CI, confidence interval; OR, odds ratio; SORT, symptom-onset to remdesivir treatment.

**Table 3** shows characteristics and outcomes of patients with SORT interval ≤9 vs >9 days. The demographic (age and gender) and clinical characteristics (comorbidities and severity distribution) were comparable between the two groups. Overall, 260 (75.1%) patients had a SORT interval of ≤9 days, while 86 (24.9%) had a SORT interval of >9 days (**Table 3**). All-cause mortality was significantly lower in the former group compared with the latter group (18.1% vs 33.7%; *P* = .004). Although not statistically significant, the median LOHS was numerically lower in patients with SORT interval ≤9 days compared with >9 days (10 v 12 days; *P* = .34).

**Table 3.** Characteristics and Outcomes of COVID-19 Patients with SORT Interval ≤9 and >9 Days.

| Characteristics | SORT Interval ≤9 days<br>(n =260) | SORT Interval >9 days<br>(n =86) | P value |
| --- | --- | --- | --- |
| Age (years), median (IQR) | 60 (49.8-69) | 59 (49.3-68) | 0.96 |
| Sex, No. (%) |  |  |  |
| Male | 202 (77.7) | 68 (79.1) | 0.91 |
| Female | 58 (22.3) | 18 (20.9) |  |
| Comorbidities, No. (%) |  |  |  |
| DM | 133 (51.2) | 40 (46.5) | 0.17 |
| HTN | 119 (45.8) | 44 (51.2) |  |
| CHD | 41 (15.8) | 13 (15.1) |  |
| CKD | 9 (3.5) | 9 (10.5) |  |
| Chronic respiratory diseases (asthma, COPD), No. (%) | 8 (3.1) | 4 (4.7) |  |
| SORT (days), median (IQR) | 5 (4-7) | 11 (10-12) | <0.001 |
| Severity, No. (%) |  |  |  |
| Moderate | 86 (33.1) | 23 (26.7) | 0.34 |
| Severe | 174 (66.9) | 63 (73.3) |  |
| Outcomes (overall), No. (%) |  |  |  |
| Discharged | 213 (81.9) | 57 (66.3) | 0.004 |
| Death | 47 (18.1) | 29 (33.7) |  |
| LOHS (days), median (IQR) | 10 (7-16) | 12 (7-17) | 0.34 |
| Mortality (disease severity), No. (%) |  |  |  |
| Moderate | 3 (1.2) | 0 (0.0) | 0.28 |
| Severe | 44 (16.9) | 29 (33.7) |  |

Abbreviations: CHD, chronic heart disease; CKD, chronic kidney disease; COPD, chronic obstructive pulmonary disease; COVID-19, coronavirus disease 2019; DM, diabetes mellitus; HTN, hypertension; IQR, interquartile range; LOHS, length of hospital stay; SORT, symptom-onset to remdesivir treatment.

In multivariate analysis, the odds of death were significantly lower in patients with SORT interval ≤9 days compared with SORT >9 days (OR = 0.44; 95% CI, 0.25-0.76; *P* = .004). When analysis was performed based on disease severity, the odds of death were significantly lower in severe COVID-19 patients with SORT interval ≤9 days compared with >9 days (OR = 0.40; 95% CI, 0.22-0.72; *P* = .003) **(Figure 1**).

AE of transaminitis was observed in 44 (12.7%) patients and acute kidney injury (AKI) in 23 (6.6%) patients. Among the 350 patients, SAEs leading to drug discontinuation were observed in 4 (1.1%) patients. Of these, 1 (0.3%) patient (moderate COVID-19) had transaminitis >5 times upper limit of normal (after 3 doses) and was discharged uneventfully; 3 (0.9%) patients had AKI (estimated glomerular filtration rate <30 mL/min after 3 doses in 2 patients and after 2 doses in 1 patient), of which 2 patients survived (moderate COVID-19, n = 1; severe COVID-19, n = 1) whereas 1 patient (severe COVID-19) died. No hypersensitivity reactions were noted.

## Discussion

This study primarily evaluated the association between SORT interval and clinical outcomes in moderate-to-severe COVID-19 and other relevant clinical endpoints, such as overall all-cause mortality, LOHS, and safety in remdesivir-treated patients. The majority of patients in this cohort had severe disease (68.5%) and reported ≥1 comorbidity (70.2%). Patients with SORT interval ≤9 days showed favorable outcomes with lower mortality (18.1% vs 33.7%) compared with SORT interval >9 days, with the difference driven largely by the severe subset. The overall all-cause mortality in this cohort was 22.0%, with higher mortality in severe disease (30.8%), particularly in mechanically ventilated patients (86.0%). Median LOHS for all patients was 11 days, slightly longer in severe disease (13 days).

Most global studies have evaluated the effects of remdesivir in COVID-19 by comparing with placebo or different durations of treatment (**Table 4**). These studies have focused primarily on clinical recovery, with no categorical reports correlating SORT interval (early vs late initiation) and mortality. Clinical benefits of early treatment have been demonstrated in rhesus macaques (a primate model) infected with SARS-CoV-2.^4^ The current hypothesis is that an anti-viral is maximally effective when administered in the earlier stage of the disease when there is active viral replication, and it is important to define that window period. Additionally, considering that the first clinical indicator of viral replication, namely symptom onset after initial exposure is an average of 5 days, minimizing the interval between symptom-onset and treatment initiation assumes greater importance. Our subgroup analysis showed a significant benefit on all-cause mortality (18.1% vs 33.7%; *P* = .004) in patients with SORT interval ≤9 days vs >9 days. Further, in those with severe disease, the odds of death were significantly lower with SORT interval ≤9 days vs >9 days (OR = 0.40; 95% CI, 0.22-0.72; *P* = .003), suggesting a beneficial effect of earlier remdesivir treatment on survival predominantly in severe disease. Reviewing prior literature, in the Adaptive Covid-19 Treatment Trial (ACTT-1) final report, median duration between symptom onset and treatment was 9 days. Patients who received remdesivir within 10 days of symptom onset had a rate ratio for clinical recovery of 1.37 (95% CI, 1.14-1.64), compared to 1.20 (95% CI, 0.94-1.52) in those who received remdesivir after 10 days; however, there was no mortality benefit mentioned.^10^ Similarly, in the study of 237 patients by Wang et al, patients treated within 10 days of symptom onset had a lower 28-day mortality vs placebo (11% vs 15%; difference −3.6% [95% CI, −16.2 to 8.9]).^7^ However, no comparison was made between early vs late treatment among remdesivir-treated patients. In the study by Goldman et al, in patients not requiring mechanical ventilation, a higher proportion who had symptoms for <10 days were discharged from the hospital than patients who had symptoms for ≥10 days before receiving the first dose of remdesivir (62% vs 49%).^9^ However, mortality with respect to SORT interval was not reported.

**Table 4.** Summary of Outcomes Reported in COVID-19 Patients Treated with Remdesivir.

| Parameters | ACTT-final report<sup>10</sup> | Wang et al, 2020<sup>7</sup> | Goldman et al, 2020<sup>9</sup> | Solidarity trial<sup>11</sup> | Current study |
| --- | --- | --- | --- | --- | --- |
| Sample size and treatment | N = 1062<br>R (n = 541) vs placebo (n = 521) | N = 237<br>R (n = 158) vs placebo (n = 78) | N = 397<br>R 5-days (n = 200) vs R 10-days (n = 197) | N = 11,266<br>R (n = 2743) vs control (n = 2708) | N = 346 |
| Baseline oxygen requirement |  |  |  |  |  |
| Room air | 75 (13.9%) vs 63 (12.1%) | 0 (0%) vs 3 (4%) | 34 (17%) vs 21 (11%) | 661 (24.1%) vs 664 (24.5%) | NIL |
| On supplemental oxygen | 232 (42.9%) vs 203 (39.0%) | 129 (82%) vs 65 (83%) | 113 (56%) vs 107 (54%) | 1828 (66.6%) vs 1811 (66.9%) | 161 (46.5%) |
| On NIV/HFNC | 95 (17.6%) vs 98 (18.8%) | 28 (18%) vs 9 (12%) | 49 (24%) vs 60 (30%) |  | 135 (39.0%) |
| On MV/ECMO | 131 (24.2%) vs 154 (29.6%) | 0 (0%) vs 1 (1%) | 4 (2%) vs 9 (5%) | 254 (9.3%) vs 233 (8.6%) | 50 (14.5%) |
| Outcomes assessed |  |  |  |  |  |
| Time to recovery, days | Median (95% CI): 10 (9-11) vs 15 (13-18) days | Median (IQR): 21.0 (13.0-28.0) vs 23.0 (15.0-28.0) days | Median (IQR): 10 (6-18) vs 11 (7-NR) days | Not reported | Not reported |
| Mortality | By day 15: 6.7% vs 11.9%<br>By day 29: 11.4% vs 15.2% | At day 28: 14% vs 13% | Day 14: 8% vs 11% | At day 28: 12.5% vs 12.7% | Overall: 22.0%<br>Moderate: 2.8%<br>Severe: 30.8% |
| LOHS, median days | Initial<sup>a</sup> median (IQR): 12 (6-28) vs 17 (8-28) | Median (IQR): 25.0 (16.0-38.0) vs 24.0 (18.0-36.0) | Median (IQR) among patients discharged on/before day 14: 7 (6-10) vs 8 (5-10) | Not reported | 11 (7-16) |
| Outcomes based on SORT |  |  |  |  |  |
| SORT<sup>b</sup>, days | Median (IQR): 9 (6-12) vs 9 (7-13) | Median (IQR): 11 (9-12) vs 10 (9-12) | Median (IQR): 8 (5-11) vs 9 (6-12) days | Not reported | Median (IQR): 6 days (4-9) |
| SORT<sup>b</sup>, groups | • ≤10 days<br>• >10 days | • ≤10 days<br>• >10 days | • <10 days<br>• ≥10 days | Not reported | • ≤9 days<br>• >9 days |
| Clinical improvement | Median time to recovery (R vs placebo):<br>• ≤10 days: 9 vs 15 (rate ratio for recovery: 1.37)<br>• >10 days: 11 vs 15 (rate ratio for recovery: 1.20) | Median (IQR) time (R vs placebo):<br>• ≤10 days: 18.0 (12.0-28.0) vs 23.0 (15.0-28.0); (hazard ratio = 1.52; 95% CI: 0.95-2.43) | Discharge rate in the overall population (<10 vs ≥10 days SORT): 62% vs 49% | Not reported | Not reported |
| All-cause mortality | Not reported | Day 28:<br>• ≤10 days: 11% vs 15%<br>• >10 days: 14% vs 10% | Not reported | Not reported | 18.1% vs 33.7% |
| LOHS, days | Not reported | Not reported | Not reported | Not reported | 10 (7-16) vs 12 (7-17) |

Abbreviations: ACTT, Adaptive Covid-19 Treatment Trial; CI, confidence interval; ECMO, extracorporeal membrane oxygenation; HFNC, high flow nasal cannula; IQR, interquartile range; LOHS, length of hospital stay; MV, mechanical ventilation; NIV, non-invasive ventilation; NR, not reported/possible to estimate; R, remdesivir; SORT, symptom onset to remdesivir treatment.
<sup>a</sup> Readmission: 5% vs 3%.
<sup>b</sup> SORT: In non-remdesivir group, it refers to time between symptom onset and receiving either placebo or non-remdesivir treatment.

The more recent WHO Solidarity trial (under review) included a large number of patients on remdesivir (n = 2743), but did not analyze outcomes based on SORT interval.^11^ The lack of benefit on mortality and other outcomes in the preliminary Solidarity trial results may be due to a ‘lumper’ phenomenon with many medications studied, and details of the relationship between SORT interval and mortality outcomes were not reported. Our study demonstrates the mortality advantage of SORT interval up to 9 days in moderate-to-severe COVID-19 indicating the window for optimal impact of the medication in this severity class. Thus, the cardinal difference between our study and other robust trials is the mortality benefit observed with earlier initiation of the drug, thereby defining the ‘SORT window.’ Though previous studies have considered 8-11 days as the median time from symptom-onset to treatment for analyzing various outcomes,^7,9,10^ our study provides an objective cut-off of 9 days for mortality benefit, and also reiterates the need to consider ‘symptom-onset’ and not ‘date of diagnosis’ as the target for intervention.

The mortality and recovery time/discharge rates reported in the published studies vary widely based on factors such as patient selection, disease severity (defined differently in various studies), treatment duration, and the stage of the pandemic reflecting change in therapeutic regimens. In our study, 76 (22.0%) patients died, a higher mortality rate compared with patients treated with remdesivir in the pivotal ACTT-1 trial (overall mortality: 6.7% at day 15; 11.4% at day 29),^10^ study by Wang et al (overall mortality: 14% at day 28),^7^ study by Goldman et al (overall mortality at day 14: 8% in 5-day and 11% in 10-day remdesivir treatment group respectively),^9^ and the Solidarity trial (overall mortality at day 28: 12.5%).^11^ The higher mortality rate in our cohort could be due to greater severity of illness, as we included moderate-to-severe cohort. While 13.9% of patients on remdesivir in the ACTT-1 and 24.1% in the Solidarity trial did not receive supplemental oxygen,^10,11^ all patients in our study required oxygen.

Our study as compared to ACTT-1,^10^ had a higher percentage of males (78.0% vs 65.1%). Patients with hypertension (47.1%) in our study were comparable to the remdesivir group in the ACTT-1 (50.6%)^10^ and the study by Wang et al (46%).^7^ However, patients with diabetes mellitus in the current study (50.0%) were higher than the ACCT-1 trial (30.8%) and the study by Wang et al (25%). Of note, COVID-19 mortality is higher in patients with these comorbidities^14,15^ and the role of remdesivir assumes greater importance. The higher percentage of males, those with diabetes mellitus, and all patients requiring oxygen are all factors which may explain the overall higher mortality in our cohort.

In the present study, the overall median (IQR) LOHS was 11 (7-16) days (moderate disease: 9 [6-12] days; severe disease: 13 [8-18] days). The median LOHS in the severe group in our study was substantially lower than in Wang et al (median [IQR]: 25.0 [16.0-38.0] days).^7^ However, comparison between the two studies is limited because of the differences in study design (retrospective vs randomized control trial [RCT]), geographical location, and local country protocols for discharge at that time in the pandemic, which may have impacted duration of hospitalization.

In our study, AKI was observed in 6.6% of patients (n = 23), lower than that reported by Antinori et al (22.8%).^16^ AE of transaminitis was observed in 44 (12.7%) patients, lower than that reported by Spinner et al (alanine aminotransferase increase: 32%-34%; aspartate aminotransferase increase: 32%).^8^ SAEs leading to drug discontinuation were observed in 4 out of 350 patients (3 [0.9%] due to AKI and one [0.3%] due to transaminitis). Our low discontinuation rates reiterate the importance of liver and renal function monitoring while on remdesivir and the adequate hydration followed in this cohort.

It is noteworthy to mention the recent studies and events that have preceded our study. Studies on several aspects of COVID-19 therapeutics, due to rapidly evolving data, have been confounded by a temporal bias of heterogeneity of treatment that can have a substantial impact on the conclusions. For example, steroids in moderate doses are being uniformly used only after July 2020, when results of the Randomized Evaluation of Covid-19 Therapy (RECOVERY) trial were released.^17^ Trials without steroids or a mixed population with/without steroids, would likely report a different outcome compared to studies with uniform steroid use. In addition, the inpatient pandemic-readiness from the point-of-view of infrastructure-personnel (e.g., intensive care unit [ICU], ventilators, high flow nasal cannula, non-invasive ventilation) is better in the latter half of the pandemic (post-June 2020) compared to earlier (March to June 2020) that may also influence outcomes. In the earlier part of the pandemic, data collection and reporting were also challenging as healthcare systems scrambled to manage the unprecedented burden, which is reflected in the minimalistic requirements of information in the Solidarity trial. Our study reflects real-world practice in the latter part of the pandemic, and includes a uniform steroid use, a well-prepared healthcare system and ICU infrastructure, and adequate access and uniform use of remdesivir. The importance of early remdesivir initiation (SORT interval ≤9 days) in our study with a clear outcome benefit indicates an impact of the medication even with steroids and ancillary therapies in moderate-to-severe COVID-19.

Our study has some limitations, such as the retrospective nature of the study and the lack of a control group. However, the main outcome measure here is the impact of SORT interval ≤9 days and >9 days, when all patients received remdesivir. On a real-world practice front, with an EUA from the FDA for remdesivir for moderate-to-severe COVID-19 at the time of the study, it was not ethically possible to do a RCT with a placebo group in this subset of sick patients. Further, our study did not evaluate the optimal duration of remdesivir treatment (5 vs 10 days), and cannot comment on the specific subset of mechanically ventilated patients, where role of the drug is controversial. Additionally, it is difficult to delineate the impact of remdesivir from other COVID-19-validated medications such as steroids, as this is one of the first studies to report uniform use of both these agents. However, the observation that mortality increased after 9 days, when steroids are known to be maximally useful, strongly suggests that the mortality benefit of SORT interval ≤9 days can be attributed to remdesivir. Additionally, we used remdesivir preparations from three different companies as the demand increased exponentially and the pharmaceutical industry geared up to handle the need.

## Conclusions

In summary, our study is one of the first to show that in the subset of hospitalized moderate-to-severe COVID-19 patients, SORT interval ≤9 days is associated with a mortality benefit. Our findings reinforce the need to consider earlier remdesivir initiation from symptom onset in moderate-to-severe COVID-19 infection.

## Supporting information

Supplemental Figure eFigure 1

STROBE checklist

## Data Availability

The de-identified data that support the findings of this study are available from the corresponding author (RMM) upon reasonable request.

## Additional information

All authors contributed to study design, data collection, and interpretation. All authors critically reviewed and approved the final manuscript for publication. All authors had full access to the complete study data and take responsibility for the integrity of the data and the accuracy of the data analysis.

## Previous Presentation

None.

## Acknowledgments

The authors acknowledge Aswin Sankar, MSc, and Pattabhi Machiraju, PhD, from Indegene Pvt. Ltd., for statistical analysis; Sudha Korwar, PhD, and Ramu Periyasamy, PhD, from Indegene Pvt. Ltd., for medical writing assistance; Michael Cutaia, MD, Pulmonary/Critical Care, Brooklyn NY, for manuscript review.

## References

1. World Health Organization. Technical guidance. Naming the coronavirus disease (COVID-19) and the virus that causes it. Accessed October 2, 2020. https://www.who.int/emergencies/diseases/novel-coronavirus-2019/technical-guidance/naming-the-coronavirus-disease-(covid-2019)-and-the-virus-that-causes-it.

2. World Health Organization. WHO Coronavirus Disease (COVID-19) Dashboard. Accessed November 2, 2020. https://covid19.who.int/.

3. Pizzorno A, Padey B, Dubois J, et al. In vitro evaluation of antiviral activity of single and combined repurposable drugs against SARS-CoV-2. Antiviral Res. 2020;181:104878. doi:10.1016/j.antiviral.2020.104878

4. Williamson BN, Feldmann F, Schwarz B, et al. Clinical benefit of remdesivir in rhesus macaques infected with SARS-CoV-2. Nature. 2020;585(7824):273–276. doi:10.1038/s41586-020-2423-5

5. US FDA. Coronavirus (COVID-19) update: FDA issues emergency use authorization for potential COVID-19 treatment. Published May 1, 2020. Accessed October 2, 2020. https://www.fda.gov/news-events/press-announcements/coronavirus-covid-19-update-fda-issues-emergency-use-authorization-potential-covid-19-treatment.

6. US FDA. FDA Approves First Treatment for COVID-19. Published October 22, 2020. Accessed October 28, 2020. https://www.fda.gov/news-events/press-announcements/fda-approves-first-treatment-covid-19.

7. Wang Y, Zhang D, Du G, et al. Remdesivir in adults with severe COVID-19: a randomised, double-blind, placebo-controlled, multicentre trial. Lancet. 2020;395(10236):1569–1578. doi:10.1016/S0140-6736(20)31022-9

8. Spinner CD, Gottlieb RL, Criner GJ, et al. Effect of Remdesivir vs Standard Care on Clinical Status at 11 Days in Patients With Moderate COVID-19. JAMA. 2020;324(11):1048. doi:10.1001/jama.2020.16349

9. Goldman JD, Lye DCB, Hui DS, et al. Remdesivir for 5 or 10 Days in Patients with Severe Covid-19. N Engl J Med. May 2020:NEJMoa2015301. doi:10.1056/NEJMoa2015301

10. Beigel JH, Tomashek KM, Dodd LE, et al. Remdesivir for the Treatment of Covid-19 — Final Report. N Engl J Med. May 2020:NEJMoa2007764. doi:10.1056/NEJMoa2007764

11. WHO Solidarity Trial Consortium. Repurposed Antiviral Drugs for COVID-19; Interim WHO SOLIDARITY Trial Results. https://www.medrxiv.org/content/10.1101/2020.10.15.20209817v1.

12. Government of India Ministry of Health and Family Welfare Directorate General of Health Services (EMR Division). CLINICAL MANAGEMENT PROTOCOL: COVID-19. Accessed October 5, 2020. https://www.mohfw.gov.in/pdf/ClinicalManagementProtocolforCOVID19.pdf.

13. World Health Organization. Clinical management of COVID-19. Interim guidance. Published 27 May 2020. Accessed 4 November 2020. https://www.who.int/publications/i/item/clinical-management-of-covid-19.

14. Cummings MJ, Baldwin MR, Abrams D, et al. Epidemiology, clinical course, and outcomes of critically ill adults with COVID-19 in New York City: a prospective cohort study. Lancet. 2020;395(10239):1763–1770. doi:10.1016/S0140-6736(20)31189-2

15. Wu C, Chen X, Cai Y, et al. Risk Factors Associated With Acute Respiratory Distress Syndrome and Death in Patients With Coronavirus Disease 2019 Pneumonia in Wuhan, China. JAMA Intern Med. 2020;180(7):934. doi:10.1001/jamainternmed.2020.0994

16. Antinori S, Cossu MV, Ridolfo AL, et al. Compassionate remdesivir treatment of severe Covid-19 pneumonia in intensive care unit (ICU) and Non-ICU patients: Clinical outcome and differences in post-treatment hospitalisation status. Pharmacol Res. 2020;158:104899. doi:10.1016/j.phrs.2020.104899

17. Dexamethasone in Hospitalized Patients with Covid-19 — Preliminary Report. N Engl J Med. July 2020:NEJMoa2021436. doi:10.1056/NEJMoa2021436

