## Supplemental Figure eFigure 1 for "A shorter symptom-onset to remdesivir treatment (SORT) interval is associated with a lower mortality in moderate-to-severe COVID-19: A real-world analysis"

**Supplementary information**

**eFigure 1. Kaplan-Meier Curve**

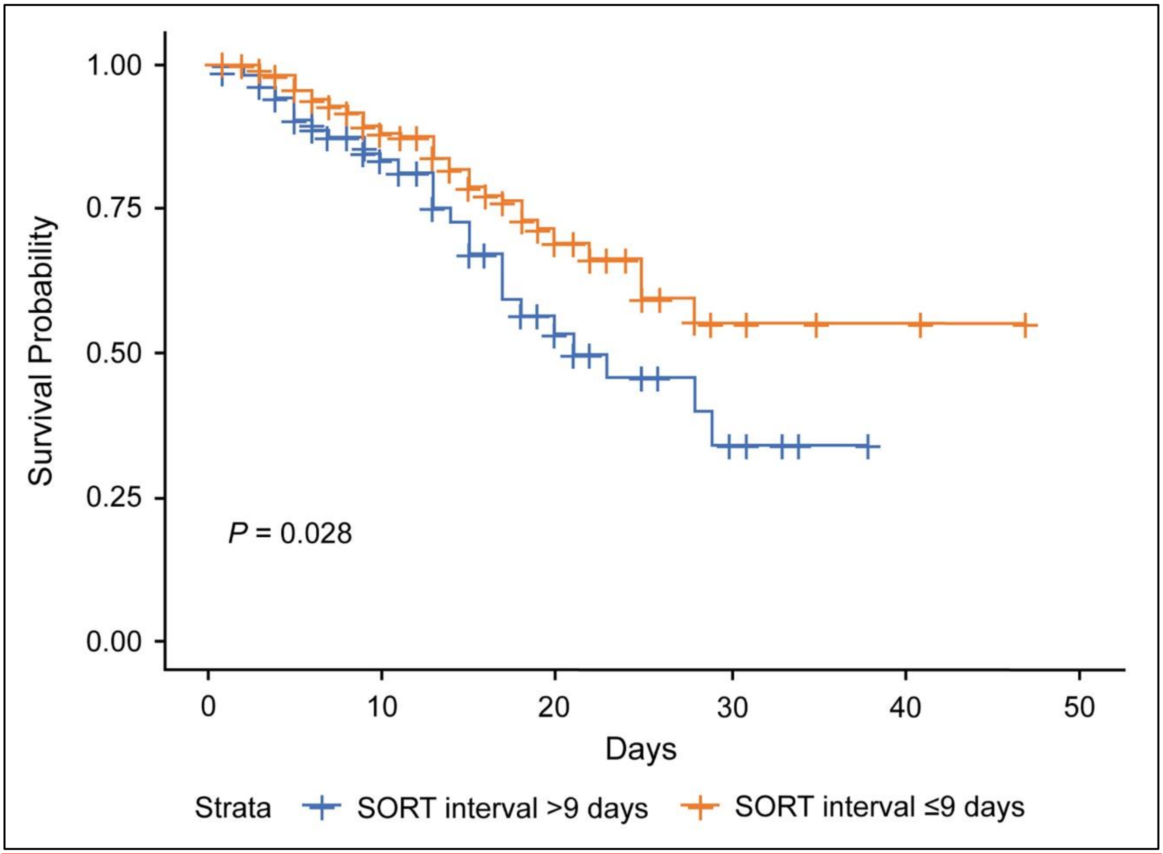


SORT, symptom onset to remdesivir treatment.
